# The Relationship between Alcohol Use Disorder, Measures of Cognitive Decline, and Alzheimer Disease Biomarkers in Older Adults

**DOI:** 10.1101/2025.05.30.25328571

**Authors:** Lisa A. Hayibor, Andrey Anokhin, Sherri Fisher, Alison Goate, Tatiana M. Foroud, Suzanne E. Schindler, Laura J. Bierut, Sarah M. Hartz

## Abstract

**Background:** Alcohol use disorder (AUD) is associated with increased risks of neuropsychiatric conditions and dementia. However, the association between Alzheimer disease (AD) and AUD is poorly characterized. To address this, we studied associations between AUD, cognition, and measures of AD neuropathology.

**Methods:** We measured a lifetime history of AUD, cognitive decline and blood biomarkers for AD (Amyloid Positivity Score 2, Aβ42/Aβ40 and p-tau217/np-tau217 ratio) in older participants from the St. Louis site of the Collaborative Study on the Genetics of Alcoholism (COGA). AUD was defined as having four or more DSM-5 AUD symptoms at the time of heaviest lifetime consumption, as most AUD related morbidity and mortality are associated with moderate/severe AUD. Cognitive decline was measured using the AD8 score (N=356), and AD biomarkers were derived from plasma measurements (N=138). We used Poisson regression models to evaluate the relationship between AUD, age, and cognitive decline, with age modeled as a piecewise linear variable by decade. Linear regressions were used to assess the association between AD blood biomarkers and AUD.

**Results:** Analyses revealed a significant association between AUD and increased cognitive decline in older adults (RR=1.31, p<0.001). No statistical association was seen between plasma AD biomarkers and AUD.

**Conclusions:** These results underscore the importance of addressing AUD as a potentially modifiable risk factor for cognitive decline in older adults. Further study is needed to understand the link between AUD and AD biomarkers.

## Introduction

Alcohol use disorder (AUD) significantly contributes to mental, physical, and cognitive disabilities, as well as preventable deaths in the US, especially among individuals aged 40 and older (Paul et al., 2024). Excessive alcohol use is estimated to contribute to over 178,000 deaths annually, making it one of the leading preventable causes of death in the US (Barr et al., 2023). Because of high levels of alcohol use in the aging population, alcohol-related injuries and diseases in older adults are expected to increase dramatically (White et al., 2023). Of particular concern are adverse effects AUD has on the brain, which may exacerbate cognitive decline and other neuropsychiatric conditions (Li et al., 2024).

Although the association between dementia and AUD has been described for centuries (Eckardt and Martin, 1986, Rehm et al., 2019), a growing body of evidence is emerging that dementia risk is increased with both AUD and heavy alcohol use (defined as >14 drinks weekly) such that excessive alcohol consumption is now considered a modifiable risk factor for dementia (Livingston et al., 2020). A 5-year longitudinal study of over 31 million people admitted to the hospital in France found that AUD was associated with a three-fold increased risk of dementia, and accounted for 56% of all dementia diagnoses under the age of 65 (Schwarzinger et al., 2018). In addition, the UK Whitehall Study, which surveyed 9,087 participants aged 35-55 and followed them for 23 years, found that heavy alcohol use was associated with a 17% increased risk of dementia relative to individuals who drank less than 10 drinks per week (Sabia et al., 2018). Despite this growing body of evidence, the precise impact of AUD on cognitive decline in older adults remains unclear.

The most common cause of dementia in the US is Alzheimer disease (AD), characterized by a slow progressive decline in cognitive function (Morris et al., 2014, Sosa-Ortiz et al., 2012). Based on our current understanding of the neuropathological processes, AD is conceptualized as a continuum in which the AD neuropathology builds up over decades, eventually resulting in the cognitive decline that defines symptomatic AD dementia (Morris et al., 2014). The neuropathological changes in AD are defined by accumulation of extracellular beta-amyloid plaques, followed by accumulation of intraneuronal tau tangles. The accumulation of these protein aggregates is accompanied by gliosis (microgliosis and astrocytosis) and neuronal loss. Because the neuropathology of AD starts approximately 20 years before the onset of clinical symptoms of AD (Bateman et al., 2012, Villemagne et al., 2013), AD has been divided into three disease stages: (1) preclinical AD; (2) a transition period where some mild cognitive impairment is evident without meeting criteria for dementia; and (3) AD dementia. The etiological mechanisms underlying these neuropathological changes remain unclear but are probably caused by both environmental and genetic factors (Bateman et al., 2012, Dubois et al., 2016). There is great interest in preclinical AD, because it represents a period when individuals at increased risk of AD dementia can be identified for preventative interventions (Dubois et al., 2016).

While AD biomarkers (amyloid and tau) can be detected through PET scans and cerebrospinal fluid (CSF) analysis, (Angioni et al., 2022, Leuzy et al., 2025), recent advances, including FDA approval, have made blood-based biomarker tests more accessible (Angioni et al., 2022, Hampel et al., 2023, Hampel et al., 2018, Hansson et al., 2022, Kiddle et al., 2018, Quest, 2023, Schindler and Bateman, 2021, Teunissen et al., 2022, U.S. Food and Drug Administration, 2025). These biomarkers offer valuable insights into early disease mechanisms, yet the role of AUD in accelerating these neuropathological changes remains poorly understood.

Despite increasing evidence linking AUD to both cognitive decline and neurodegeneration, significant gaps remain in understanding how AUD influences cognitive trajectories in aging adults. Most prior studies have focused on dementia risk rather than early cognitive decline in midlife, and few have examined whether AUD contributes to earlier biomarker changes associated with AD pathology. To address these gaps, we investigated the relationship between AUD, cognitive decline and AD biomarkers. Because most of the morbidity and mortality related to AUD are associated with moderate and severe AUD, for this study we defined AUD as those who meet DSM-5 criteria for moderate or severe AUD (4 or more symptoms of AUD). Likewise, participants without AUD refers to those who do not meet the criteria for moderate or severe AUD, including individuals with mild AUD (2-3 DSM-5 symptoms), and those with 0-1 DSM-5 symptoms.

### Materials and Methods

#### Participants

The Collaborative Study on the Genetics of Alcoholism (COGA) is a family-based longitudinal study that was initiated in 1989 and has since continued to follow participants with AUD and community-based comparison participants and their families.(COGA, 2022). For the current study, we selected COGA participants at Washington University in St. Louis who were born before 1974, had completed a previous Semi-Structured Assessment for the Genetics of Alcoholism (SSAGA) interview, had a prior electroencephalogram (EEG), and had genetic data available. In addition to completing another SSAGA interview, participants in the current study were also asked to complete a cognitive assessment and donate a blood sample for AD biomarker testing. Our analysis sample consisted of 413 participants who completed a SSAGA interview and who either: 1) had data available from the SIST (n=356); or 2) contributed a blood sample for AD biomarker testing (n=138). This study was approved by the Washington University Institutional Review Board, and all participants gave informed consent.

#### Cognitive assessment

Cognitive function was assessed using the Structured Interview and Scoring Tool- Massachusetts Alzheimer’s Disease Research Center (SIST) (Okereke et al., 2012). The SIST includes measures of cognitive impairment and functional decline, incorporating structured questions that assess memory, executive function, and activities of daily living, and was administered to participants by trained research assistants. For the current analysis, the Eight-item Informant Interview to Differentiate Aging and Dementia (AD8) (Galvin et al., 2007, Galvin et al., 2005, Galvin et al., 2006), a widely used informant-based screening tool for cognitive impairment, was generated using responses from the SIST assessment to evaluate early signs of cognitive decline. Scores range from 0 to 8 with higher AD8 scores indicating greater cognitive impairment, and a cutoff score of >1 suggesting significant functional decline. The mapping of SIST items to AD8 scoring is given in Supplementary Table 1.

#### Alcohol Use Disorder

We assessed lifetime history of AUD as part of the SSAGA, a structured diagnostic interview designed to assess a wide range of psychiatric and substance use disorders based on DSM criteria (Bucholz et al., 1994). Because moderate to severe AUD accounts for the majority of alcohol-related morbidity and mortality, we defined AUD in this study as having four or more DSM-5 AUD symptoms at the time of their heaviest lifetime consumption (American Psychiatric Association, 2013). Participants who did not meet these criteria for moderate or severe AUD, including those with mild AUD (2-3 symptoms) or no AUD (0-1 symptoms) were classified as without AUD.

#### AD biomarker testing

Participants’ blood samples were collected and processed into plasma on site, and 0.5 mL plasma aliquots were transferred into polypropylene cryovials that were frozen (−80°C) within 2 hours of phlebotomy. The resulting plasma samples were shipped on dry ice to C2N Diagnostics (St. Louis, Missouri) for processing and analysis. C2N Diagnostics used their LC-MS/MS analytical platform to quantify plasma Aβ42, Aβ40, p-tau217, np-tau217 concentrations (all pg/mL), and calculated the plasma Aβ42/Aβ40, and p- tau217/np-tau217 concentration ratios as previously described (Hu et al., 2022, Meyer et al., 2024, West et al., 2021). Amyloid Positivity Score 2 (APS2), a function of Aβ42/Aβ40 and p-tau217/np-tau217 was calculated as part of the PrecivityAD2 test, as previously described (Meyer et al., 2024).

#### Statistical Analyses

All statistical analyses were performed using SAS 9.4 (SAS Institute Inc., 2024) or R version 4.3.2.(R Core Team, 2023). Descriptive statistics were reported for the study, with t-tests used for continuous and ordinal variables and chi-square tests for categorical variables. Gender was analyzed as a categorical variable (male, female) and AUD was analyzed as a dichotomized variable (AUD, without AUD). Missing data were excluded from the analyses to ensure the robustness of the results. Statistical significance was defined at an alpha level of 0.05.

The primary outcome variable for cognitive decline, AD8, is an ordinal variable consisting of 9 levels measuring cognitive impairment and therefore Poisson regression was used to evaluate the association of AD8 score with AUD and age. AD8 score was coded as 1 to 9 to meet the requirement of Poisson regression, which assumes ordinal outcomes start at 1. Because the effect of age on cognitive measures varies over time (Salthouse, 2012), age was modeled as piecewise linear by decade (i.e. linearity was assumed within each decade but slopes were allowed to vary between ages 50-59, 60-69, and 70 or more). The AD biomarkers APS2 and Aβ42/Aβ40 were modeled as continuous variables using linear regression. Censored regression was used to model p-tau217 ratio due to a number of participants having values below the level of detection.

## Results

The demographic characteristics of the study participants are outlined in Table 1. AUD was more prevalent in the younger group (<65 years) with 115 participants (44%) reporting a lifetime diagnosis of AUD compared to 47 (31%) in the older group (≥65 years). While gender distribution was balanced in the younger AUD group, older adults with AUD were predominantly male (70%). In both age groups, the majority of participants were White, regardless of AUD status.

**Table 1.** Demographic Characteristics of COGA Participants by Age and Alcohol Use Disorder (AUD) status

|  | Overall | Age <65 (N=260) |  | Age ≥65 (N=153) |  |
| --- | --- | --- | --- | --- | --- |
|  |  | with AUD | without AUD | with AUD | without AUD |
| **N** | 413 | 115 | 145 | 47 | 106 |
| **Age** |  |  |  |  |  |
| Mean (SD) | 62.8 (8.4) | 57.4 (4.8) | 57.8 (4.7) | 69.4 (4.0) | 72.6 (5.6) |
| **Gender n (%)** |  |  |  |  |  |
| Female | 237 (57) | 56 (49) | 100 (69) | 14 (30) | 67 (63) |
| Male | 172 (42) | 56 (49) | 44 (30) | 33 (70) | 39 (37) |
| Missing | 4 (1) | 3 (3) | 1 (1) | 0 (0) | 0 (0) |
| **Race n (%)** |  |  |  |  |  |
| Black | 70 (17) | 13 (11) | 40 (28) | 10 (21) | 7 (7) |
| Other race | 3 (1) | 1 (1) | 2 (1) | 0 (0) | 0 (0) |
| White | 340 (82) | 101 (88) | 103 (71) | 37 (79) | 99 (93) |
| **AD8 Score** |  |  |  |  |  |
| n | 356 | 81 | 140 | 29 | 106 |
| Mean (SD) | 2.4 (2.0) | 2.7 (2.2) | 2.0 (1.8) | 3.6 (2.3) | 2.5 (1.9) |
| AD8 >1 n (%) | 211 (59) | 52 (64) | 71 (51) | 22 (76) | 66 (62) |
| **APS2** |  |  |  |  |  |
| n | 138 | 32 | 42 | 16 | 48 |
| Mean (SD) | 22.7 (21.3) | 15.9 (9.7) | 14.1 (12.0) | 28.8 (25.1) | 32.8 (26.9) |
| APS2 >14.5 n(%) | 70 (51) | 12 (38) | 12 (29) | 9 (56) | 37 (77) |
| APS2 >47.5 n(%) | 15 (11) | 0 (0) | 1 (2) | 2 (13) | 12 (25) |
| **Aβ42/Aβ40** |  |  |  |  |  |
| n | 138 | 32 | 42 | 16 | 48 |
| Mean (SD) | 0.1 (0.0) | 0.1 (0.0) | 0.1 (0.0) | 0.1 (0.0) | 0.1 (0.0) |
| AB42/40 <0.1 n(%) | 89 (65) | 21 (66) | 23 (55) | 12 (75) | 33 (68) |
| **p-tau 217/np-tau217** |  |  |  |  |  |
| n | 138 | 32 | 42 | 16 | 48 |
| Mean (SD) | 3.1 (3.7) | 2 (1.1) | 2 (1.1) | 4.1 (6.1) | 4.3 (4.7) |
| p-tau 217 ratio>1.55 n(%) | 87 (63) | 16 (50) | 20 (48) | 12 (75) | 39 (81) |
| p-tau 217 ratio>4.2 n(%) | 20 (15) | 2 (6) | 1 (2) | 2 (13) | 15 (31) |

There were significant differences in AD8 scores between participants with and without AUD. The mean AD8 score was higher in participants with AUD across both age groups (2.7 for <65, 3.6 for >=65) compared to those without AUD (2.0 for <65, 2.5 for >=65). Similarly, there was a higher proportion of participants with an AD8 score >1 among AUD participants (64% for <65, 76% for >=65) compared to the non- AUD group (51% for <65, 62% for >=65). Despite differences in cognitive scores, no statistical differences were seen in the biomarker levels based on AUD for either age group.

We investigated whether AUD is associated with increased risk of cognitive decline, as measured by AD8. The association of AD8 score with age, and AUD was estimated using Poisson regression and is shown in Figure 1. As expected, AD8 scores gradually increased with age, with the slope varying over time. After adjusting for age, those with AUD were expected to score 1.3 times higher on the AD8 than those without AUD (95% CI 1.2-1.5, p<0.001). No statistical interaction was seen between AUD and age with respect to estimated AD8 score.

**Figure 1.**
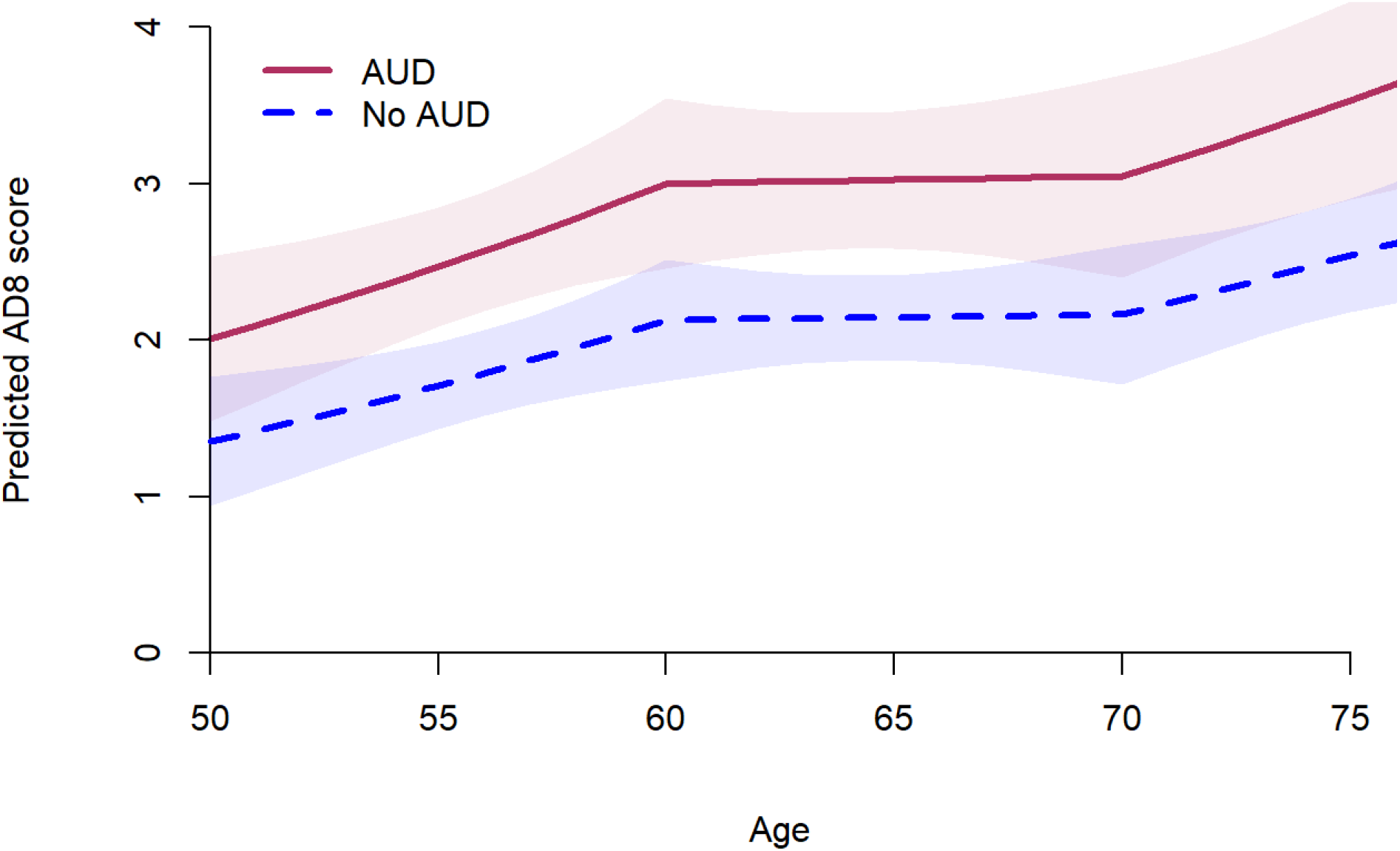
Expected AD8 score by age, for those with alcohol use disorder (AUD) and those without AUD. Computations are from Poisson regression; estimated AUD RR=1.31, 95% CI 1.16-1.48, p<0.001.

We next investigated whether individuals with AUD had evidence of increased brain amyloid deposition relative to those without AUD. We measured two plasma biomarkers that are strongly associated with amyloid deposition in the brain as measured by amyloid PET: p-tau 217/np-tau217 ratio and Aβ42/Aβ40. These two biomarkers are combined to create a third biomarker: APS2. We plotted the biomarkers as a function of age stratified by AUD diagnosis (Figure 2). For each biomarker, blue lines indicate previously estimated cutoffs for amyloid positivity based on amyloid PET centiloid (CL) values in different populations: the dashed blue line indicates the optimal cutpoint for distinguishing PET Centiloid >20 among 1080 cognitively unimpaired participants (Rissman et al., 2024); the solid blue line indicates the optimal cutpoint for distinguishing PET CL >25 among N=583 cognitively impaired participants (Meyer et al., 2024). Higher levels of brain amyloid deposition are associated with elevated APS2 and p-tau217/np-tau217 ratios, but lower Aβ42/Aβ40 values. For all three of these related biomarkers, AUD was associated with levels corresponding to more elevated brain amyloid deposition, but this did not reach statistical significance.

**Figure 2:**
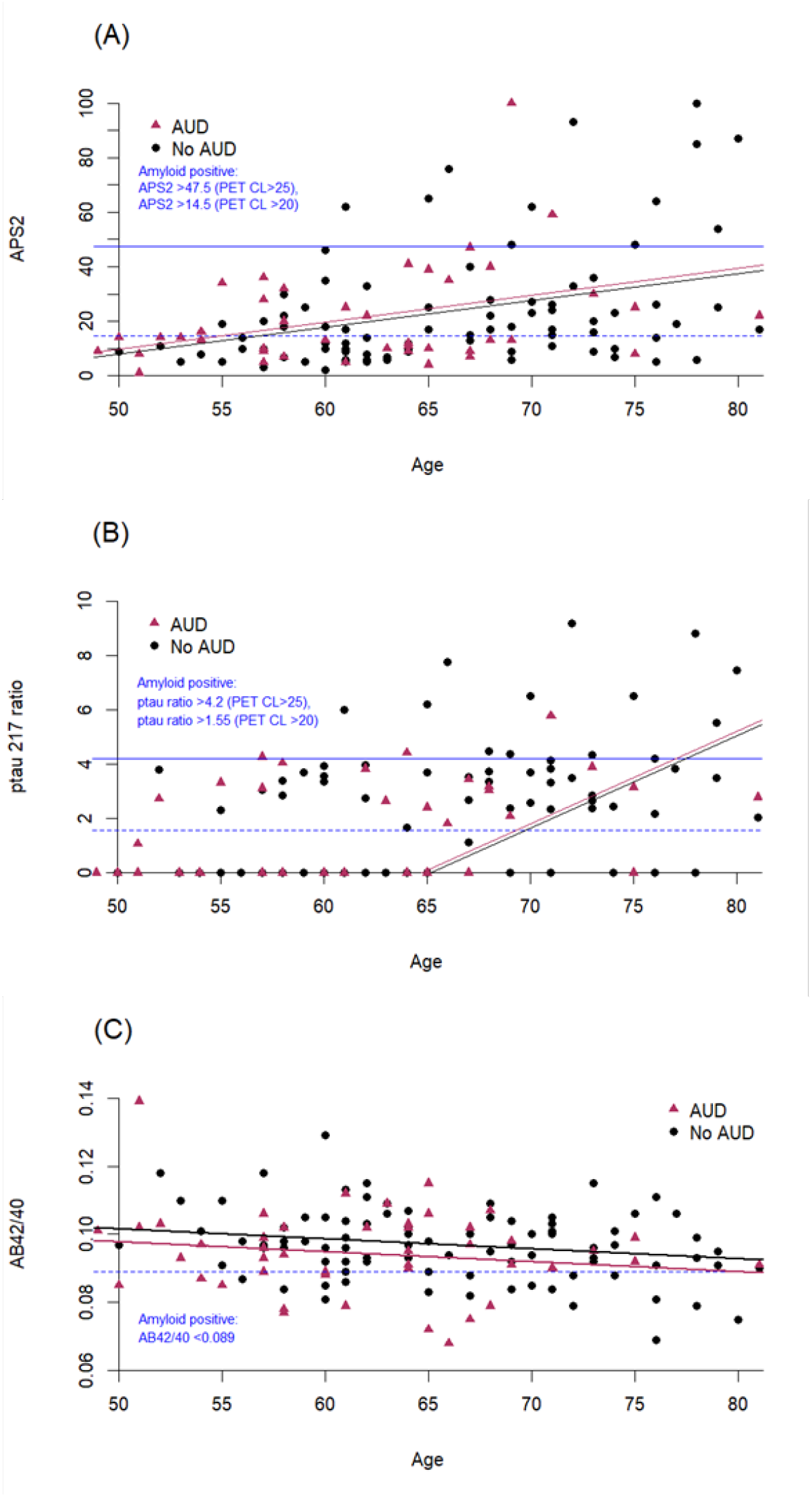
Plots of age relative to Alzheimer disease (AD) biomarkers: APS2 (A), p-tau217/np-tau217 (B), and Aβ42/Aβ40 (C). Black and red lines correspond to regression lines for biomarker ∼ age for AUD (red) and no AUD (black). Although the estimated biomarker levels correspond to increased amyloid deposition for all biomarkers, no statistical difference between the two lines is seen (p>0.05). Blue lines indicate cutoffs for amyloid positivity (dashed line is cutoff for PET CL >20, solid line is cutoff for PET CL >25).

## Discussion

Alcohol use disorder (AUD) has been extensively linked to adverse health outcomes, including cognitive impairment and neurodegeneration, particularly in aging populations(Li et al., 2024, Paul et al., 2024). Our findings provide further evidence supporting the association between AUD and cognitive decline, as measured by AD8 scores. With increasing age, participants with AUD exhibited higher AD8 scores than their non-AUD counterparts. Additionally, plasma AD biomarker findings suggest that AUD may increase brain amyloid deposition, though the association was not statistically significant. These findings highlight AUD as a risk factor for cognitive decline among older adults, reinforcing the importance of early detection and intervention to mitigate long-term neurological consequences.

Our study aligns with prior research suggesting that AUD is a modifiable risk factor for dementia, particularly Alzheimer disease (AD) (Livingston et al., 2020). Previous large-scale cohort studies, such as the French longitudinal study of over 31 million hospital patients (Schwarzinger et al., 2018) and the UK Whitehall Study (Sabia et al., 2018), have demonstrated a strong association between excessive alcohol consumption and an increased risk of dementia. We expanded upon these studies by showing that cognitive decline associated with AUD can be detected even before clinical dementia manifests. This supports the notion that AUD-related neurodegeneration may begin earlier in life and progress over time, emphasizing the need for interventions in at-risk individuals before cognitive symptoms become severe.

While the primary focus of this study was cognitive function as measured by AD8 scores, we also explored potential biological mechanisms by examining biomarkers associated with AD pathology, including amyloid-beta (Aβ42/Aβ40 ratio) and tau phosphorylated at position 217(p-tau217). Our results suggest that biomarker differences may exist between AUD and non AUD participants, although this association is not statistically significant. This finding is consistent with the hypothesis that AUD accelerates pathological processes associated with AD, potentially increasing the risk of later cognitive decline. Given that biomarker alterations were observed in aging adults with AUD, future research should explore whether early interventions targeting problematic alcohol consumption could slow or prevent cognitive decline before significant neurodegeneration occurs.

This study has several strengths, including the use of a well-characterized cohort, rigorous cognitive assessments, and biomarker analyses. However, there are limitations to consider. While AD8 is a validated cognitive screening tool, it does not provide a full clinical diagnosis of dementia, and future studies should incorporate comprehensive neuropsychological assessments. Additionally, the study’s sample size for biomarker analyses was relatively small, particularly in older AUD groups, which may have limited statistical power to detect subtle differences. Despite this, the specific findings of this study remain important, as they provide valuable insights into the relationship between AUD, cognitive decline and AD biomarkers in older adults. These insights highlight the importance of early intervention strategies to reduce alcohol-related cognitive impairment. Future studies should focus on longitudinal investigations with larger samples to further clarify the relationship between AUD, cognitive decline, and AD pathology, as well as strategies to mitigate the long-term neurological effects of AUD.

## Data Availability

All data produced in the present study are available upon request

https://cogastudy.org/resources-for-researchers/#accessing-coga-data

