## Supplementary Table 1 for "The Relationship between Alcohol Use Disorder, Measures of Cognitive Decline, and Alzheimer Disease Biomarkers in Older Adults"

**Supplementary Table 1:** SIST items used for AD8 estimation.

| **AD 8 item** | **SIST item(s) and definition for positivity**  Compared to 5-10 years ago, do you ... |
| --- | --- |
| 1. Problems with judgment (e.g., problems making decisions, bad financial decisions, problems with thinking) | Have more difficulty handling everyday problems? (“Slightly”/”Increasingly”)  ***or*** Rely on others to figure out problems or make decisions? (“Somewhat more”/ “A lot more”)  ***or*** Get confused by handling more than one problem/task at a time? (“Sometimes”/”Almost always”)  ***or*** Have more trouble with driving decisions such as merges, left turns, lane changes, or stopping at lights? (“A little more trouble making driving decisions”/”Clear difficulty with decisions”)  ***or*** Have close calls or driving accidents or mishaps more often? (“Yes, but probably not his/her fault”/”Yes, and probably at fault”)  ***or*** Have changes in your overall ability to apply judgment in situations, or to solve problems and make decisions? (“Good judgment, but not as good as before”/”Judgment is now only fair”) |
| 2. Less interest in hobbies/activities | Have increased difficulties at your current job or volunteer activities due to memory and/or thinking problems? (“Occasionally; sometimes need help”/ “Often; needs more supervision”)  ***or*** Have decreased responsibilities at your current job or volunteer activities due to memory and/or thinking problems? (“May decrease responsibilities/ take on less to do”/“Significant decrease in responsibilities”)  ***or*** Seem to be less active in social activities? (“Somewhat”/”Significantly”)  ***or*** Have more difficulty in handling roles and responsibilities in social activities? (“Somewhat; noticeable but mild”/”Significantly; very noticeable”)  ***or*** Deliberately reduce your involvement or level of responsibility in social activities, due to problems with memory or thinking? (“Somewhat”/”Significantly”)  ***or*** Seem less involved in your hobbies overall? (“Somewhat”/”Significantly”) |
| 3. Repeats the same things over and over (questions, stories, or statements) | Do you repeat the same questions or anecdotes over again? (“May repeat within same week”/”May repeat in same day”) |
| 4. Trouble learning how to use a tool, appliance, or gadget (e.g., VCR, computer, microwave, remote control) | Do you have more difficulty learning how to use new appliances? This would include things like computers, phones, or cooking or cleaning equipment. (“Some difficulty learning, but ultimately succeeds”/”Unable to learn”) |
| 5. Forgets correct month or year | Do you have more difficulty keeping track of the date and/or day? (“Sometimes doesn’t know the exact date”/”Often doesn’t know date/day”) |
| 6. Trouble handling complicated financial affairs (e.g., balancing checkbook, income taxes, paying bills) | Need more help managing finances, for example, organizing bills, assembling tax documents, or tracking savings or investments? (“Needs help/advice (a change for him/her)”/”No longer does independently”)  ***or*** Have any problems managing a checkbook? (“Occasional errors”/”Many errors”)  ***or*** Ever bounce any checks? (“Happens 2-3 times/ year; this is a change”/”Repeatedly”)  ***or*** Forget to pay bills, or pay the same bill twice? (“Forgets or double-pays bills 2-3 times/year”/”Repeatedly; needs reminding”)  ***or*** Have any recently-developed systems or new strategies to handle finances because of problems with thinking? (“Yes, and new strategies are working well”/”Yes, but problems persist”)  ***or*** Seem less likely to be able to do the kinds of mental tasks involved in your former primary job, because of current problems with memory or thinking? (“Current problems would cause some difficulty”/”Current problems would cause a lot of difficulty”) |
| 7. Trouble remembering appointments | Rely more on others to remember appointments? (“Somewhat more”/”Much more”)  ***or*** Depend more on a calendar to remember appointments? (“Somewhat more”/”Much more”) |
| 8. Daily problems with thinking and/or memory | Have more problems with memory or thinking? (“Significant change”) |

The total AD8 score is calculated as the sum of all eight items. Each item is scored as 1 if any corresponding SIST item is reported and 0 otherwise. Scores range from 0 to 8, with higher scores indicating increased cognitive decline.
